# Embedding community and public voices in co-created solutions to mitigate antimicrobial resistance (AMR) in Thailand using the ‘Responsive Dialogues’ public engagement framework

**DOI:** 10.1101/2023.10.06.23296658

**Authors:** Tassawan Poomchaichote, Niyada Kiatying-Angsulee, Kanpong Boonthaworn, Bhensri Naemiratch, Supanat Ruangkajorn, Ravikanya Praparsavath, Chaiwat Thirapantu, Karnjariya Sukrung, Direk Limmathurotsakul, Anne Osterrieder, Phaik Yeong Cheah

## Abstract

The use of antimicrobials in Thailand has been reported as one of the highest in the world in human and animal sectors. Our engagement project aimed to improve our understanding of the issue of antimicrobial use and antimicrobial resistance (AMR) among adult Thai communities, and to co-create locally relevant solutions to AMR, especially those focusing on raising awareness to improve related policies in Thailand.

We conducted a series of online and in-person ‘conversations’ according to Wellcome’s ‘Responsive Dialogues’ engagement approach, designed to bring together different voices to solve complex problems such as AMR. This approach enabled key AMR stakeholders and policy makers to hear directly from communities and members of the public, and vice versa. Conversations events took place between 25 November 2020 and 8 July 2022, and we engaged 179 AMR key stakeholders and members of the public across Thailand.

The issues we found were: there were quite a lot of misunderstandings around antimicrobials and AMR; participants felt that communications and engagement around antimicrobial resistance had limited reach and impact; asking for and taking antibiotics for self-limiting ailments is a social norm in Thailand; and there appeared to be a wide availability of cheap antimicrobials. To mitigate the spread of AMR, participants suggested that the messages around AMR should be tailored to the target audience, there should be more initiatives to increase general health literacy, there should be increased availability of AMR related information at the local level and there should be increased local leadership of AMR mitigation efforts.

**Thaiclinicaltrials.org registration:** TCTR20210528003 (28/05/2021)

**KEY MESSAGES:** *What is already known on this topic:*

⇒ Antimicrobial resistance (AMR) is a complex and systemic problem, which will require collaboration from many sectors. Any policies and interventions to address AMR need to be specific to the local context, ethical, and consider the affected communities.

*What this study adds:*

⇒ The regional spread and the depth of our community ‘conversations’ (e.g. 2.5-3 days for in-person ‘conversations’) enabled us to explore topics and important elements of solutions around AMR, summarise common themes, and compare views across different regions and communities in Thailand.

*How this study might affect research, practice or policy:*

⇒ Our project revealed some important ‘building blocks’ or elements for AMR-related policy, in particular relating to communications and engagement: messages around AMR should be tailored to the target audience, there should be more initiatives to increase general health literacy, there should be increased availability of AMR related information at the local level and there should be increased local leadership of AMR mitigation efforts.

## INTRODUCTION

Antimicrobial resistance (AMR) is the ability of microorganisms to stop antimicrobial drugs from working effectively against them ^1-3^. A recent study estimated that in 2019, 4.95 million deaths globally were associated with bacterial AMR, including 1.27 million deaths that were directly attributable to AMR ^4^.

AMR has been characterized as a ‘super-wicked’ problem that is systemic, multi-sectorial, multidisciplinary, borderless, global, and ethically challenging ^5-7^. To maximise success, policies and interventions aiming to mitigate the problem of AMR need to be context-specific and locally driven.

Thailand is an upper-middle income country with a population of approximately 70.2 million people ^8 9^. It has a high AMR burden ^10^ and high excess mortality due to hospital-acquired antimicrobial resistant infections ^11^. It is estimated to have one of the highest antibiotics uses in both human and animal sectors among countries that have published official data on national antibiotic consumption ^12-14^. In Thailand, by law, most oral antimicrobials, except tuberculosis drugs, can be dispensed by licensed pharmacists at pharmacies without a prescription from a qualified doctor ^15^. The general public have a limited understanding of AMR, as surveys showed ^16 17^. In some communities, antibiotics are perceived as a ‘quick fix’ for care, productivity, hygiene and social inequity ^18 19^.

In this paper we report our findings from piloting a dialogue-based public engagement framework in Thailand. Developed by Wellcome, the “Responsive Dialogues on Drug Resistant Infections” framework was designed to bring together multiple stakeholders and communities across the ‘One Health’ spectrum to tackle complex problems such as AMR right in communities that are most affected ^20^. The ‘responsive dialogue’ format enables AMR stakeholders and policy makers to hear directly from communities and members of the public and vice versa. The intended outcomes are to bridge the gap between policy and implementation, obtain a wide range of views, and facilitate sustained bi-directional interactions among stakeholders and people from various communities^20^. In 2019, Wellcome funded two pilots in Thailand and Malawi. This paper summarises the issues relevant to Thailand identified by our participants, and their co-created solutions.

The main objectives of our project were 1) to improve our understanding of the issue of antimicrobial use and AMR among adult Thai communities, and 2) to co-create locally relevant solutions to AMR, thereby improving policies for reducing the burden of AMR in Thailand ^21^. For practical reasons, we narrowed our scope to interventions and solutions that are actionable by those who participate in the project, in particular those related to engagement and awareness around AMR. This aligns with Strategy 5 (public knowledge and awareness of appropriate use of antimicrobials) of the Thailand National Strategic Plan on Antimicrobial Resistance (NSP-AMR) ^22 23^. Our project was intended to support the development of Strategy 5 of the new NSP-AMR 2023-2027.

### “AMR Dialogues” in Thailand – a multi-phased approach combining in-person and online conversation events

Our “AMR Dialogues” project was conducted in three phases, following guidance of the Wellcome Responsive Dialogues toolkit ^20^. These were: Phase I, ‘Groundwork’, which involved stakeholder mapping and ‘Planning Conversations’ with stakeholders, and inviting participants from communities; Phase II, ‘Community Conversations’, held in four regions of Thailand, as well as two virtual ‘National Conversations’; and Phase III, evaluation and feedback to stakeholders and participants (Table 1; Suppl. Table 2).

**Table 1:**
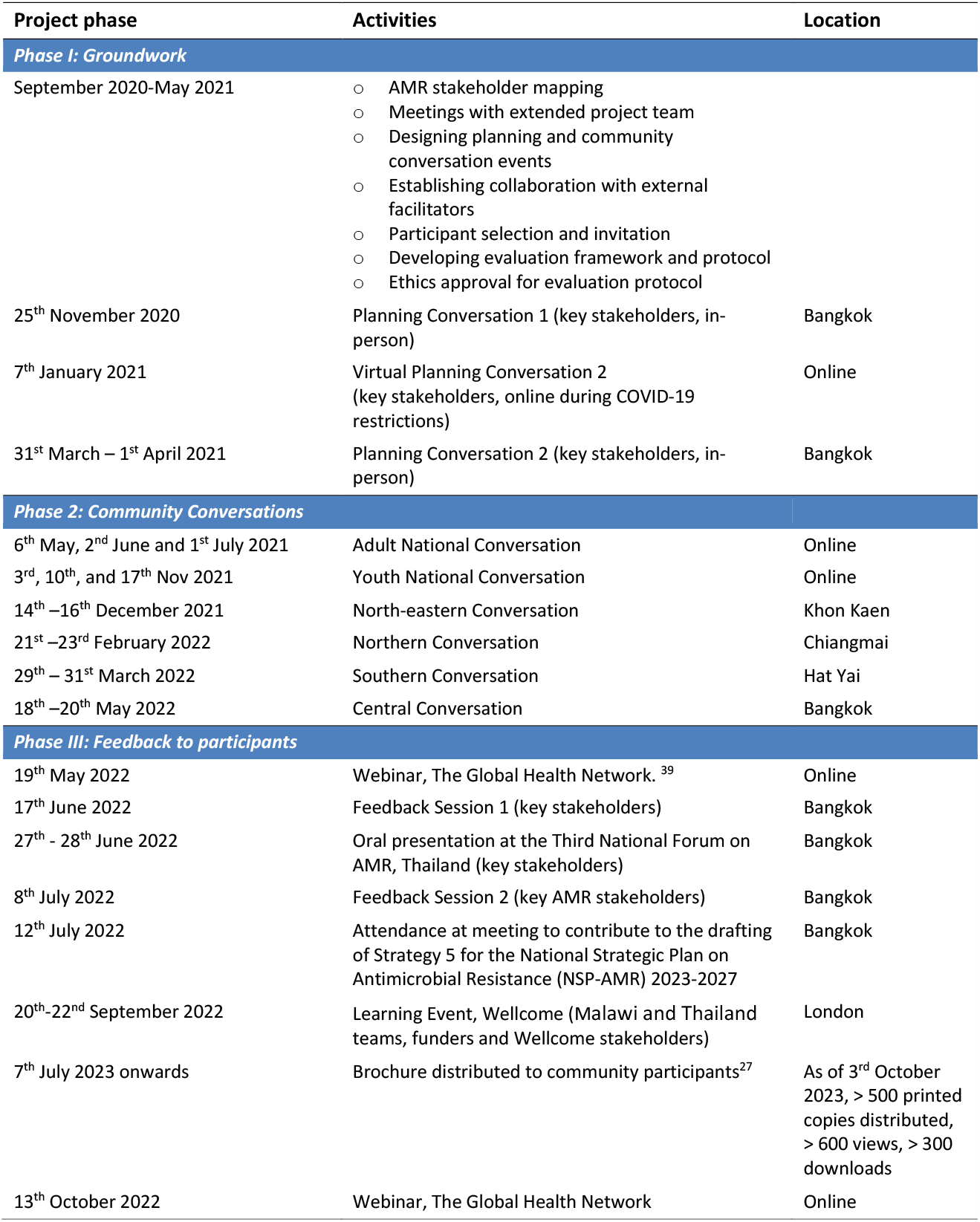
Timeline of the ‘AMR Dialogues’ project, showing the three project phases, key activities undertaken in each phase, and locations.

| Project phase | Activities | Location |
| --- | --- | --- |
| **Phase I: Groundwork** |  |  |
| September 2020-May 2021 | ○ AMR stakeholder mapping<br>○ Meetings with extended project team<br>○ Designing planning and community conversation events<br>○ Establishing collaboration with external facilitators<br>○ Participant selection and invitation<br>○ Developing evaluation framework and protocol<br>○ Ethics approval for evaluation protocol |  |
| 25<sup>th</sup> November 2020 | Planning Conversation 1 (key stakeholders, in-person) | Bangkok |
| 7<sup>th</sup> January 2021 | Virtual Planning Conversation 2 (key stakeholders, online during COVID-19 restrictions) | Online |
| 31<sup>st</sup> March – 1<sup>st</sup> April 2021 | Planning Conversation 2 (key stakeholders, in-person) | Bangkok |
| **Phase 2: Community Conversations** |  |  |
| 6<sup>th</sup> May, 2<sup>nd</sup> June and 1<sup>st</sup> July 2021 | Adult National Conversation | Online |
| 3<sup>rd</sup>, 10<sup>th</sup>, and 17<sup>th</sup> Nov 2021 | Youth National Conversation | Online |
| 14<sup>th</sup>–16<sup>th</sup> December 2021 | North-eastern Conversation | Khon Kaen |
| 21<sup>st</sup>–23<sup>rd</sup> February 2022 | Northern Conversation | Chiangmai |
| 29<sup>th</sup> – 31<sup>st</sup> March 2022 | Southern Conversation | Hat Yai |
| 18<sup>th</sup>–20<sup>th</sup> May 2022 | Central Conversation | Bangkok |
| **Phase III: Feedback to participants** |  |  |
| 19<sup>th</sup> May 2022 | Webinar, The Global Health Network. <sup>39</sup> | Online |
| 17<sup>th</sup> June 2022 | Feedback Session 1 (key stakeholders) | Bangkok |
| 27<sup>th</sup> - 28<sup>th</sup> June 2022 | Oral presentation at the Third National Forum on AMR, Thailand (key stakeholders) | Bangkok |
| 8<sup>th</sup> July 2022 | Feedback Session 2 (key AMR stakeholders) | Bangkok |
| 12<sup>th</sup> July 2022 | Attendance at meeting to contribute to the drafting of Strategy 5 for the National Strategic Plan on Antimicrobial Resistance (NSP-AMR) 2023-2027 | Bangkok |
| 20<sup>th</sup>-22<sup>nd</sup> September 2022 | Learning Event, Wellcome (Malawi and Thailand teams, funders and Wellcome stakeholders) | London |
| 7<sup>th</sup> July 2023 onwards | Brochure distributed to community participants<sup>27</sup> | As of 3<sup>rd</sup> October 2023, > 500 printed copies distributed, > 600 views, > 300 downloads |
| 13<sup>th</sup> October 2022 | Webinar, The Global Health Network | Online |

#### Phase I: Groundwork

This phase focused on understanding and mapping the ‘AMR ecosystem’ in Thailand. To produce a Thai AMR stakeholder map, we sought input from: AMR experts; existing literature ^20 24^; stakeholders attending the ‘Planning Conversations’; and members of the ‘Bangkok Health Research and Ethics Interest Group’ (an established MORU public advisory group). We identified eight major stakeholder groups (Suppl. Table 1).

As part of Phase I, we conducted three “Planning Conversations” with key AMR stakeholders: two face-to-face meetings in Bangkok (Suppl. Table 1) and one virtual workshop due to COVID-19 restrictions.

Stakeholders (defined as anyone with an interest in AMR) included government officers, researchers and experts, healthcare providers, science communication experts, and non-governmental organization representatives with an interest in AMR or related issues such as animal health and environmental issues (e.g., Greenpeace and World Animal Protection). These stakeholders were approached by personal contacts through existing networks of project team members (DL and NK), who were already involved in Thai AMR policy making and had worked in civil society for more than a decade. We specifically invited members of the Thai NSP-AMR subcommittee involved in writing the NSP-AMR 2023-2027 and previous versions to participate. The outcomes of the Planning Conversations were: to understand the AMR ecosystem in Thailand; align our project with the NSP-AMR; identify other stakeholders who should be involved; identify participants for the Community Conversations, provide input into the conversation event agenda, and map out current issues on AMR for discussion with participants. In our virtual workshop we discussed previous AMR engagement initiatives.

For public involvement, we piloted some of the activities and information materials used in Community Conversations with the Bangkok Health Research and Ethics Interest Group^25^ and revised them based on their feedback.

#### Evaluation

The evaluation process was integrated into the project from its outset to have a continuous feedback loop^21^. Evaluation included feedback forms immediately after the Conversations and focus group discussions (FGDs) with selected participants. An estimated 80% of attendees participated in the evaluation. In total, we received 266 evaluation forms; 26 attendees participated in FGDs, and 16 in in-depth interviews (IDIs). The findings informed the subsequent conversations and ensured that the objectives of each phase of the project were achieved. The protocol, information sheet and consent form for the evaluation component of the project was approved by the ethics committee of the Thailand

Institute for the Development of Human Subject Protection (IHRP2021059) and the Oxford University Tropical Research Ethics Committee (OxTREC529-21).

#### Phase II: Community Conversations

We facilitated two types of Community Conversations: in-person regional conversations and online national conversations. By using this two-pronged approach, we hoped to be as inclusive as possible and maximise the chance of having a diverse audience, to obtain a wide range of perspectives and personal experiences. Each Community Conversation was guided by the following sequence as detailed in the ‘Responsive Dialogues’ toolkit ^20^: 1) introduce AMR and explore local issues related to AMR, 2) ideate and co-create local solutions and 3) choose promising/feasible solutions to take forward.

For our Community Conversations, we invited participants according to our selection criteria described previously.^21^ Briefly, we aimed to include people from diverse backgrounds (age, gender, education levels, professions, socioeconomic backgrounds, disabilities), including those who might be affected by any changes in policy around antimicrobials, those who can influence change, and those who might be disproportionately affected by AMR and changes in related policies. We relied on our own networks, which were broad due to previous AMR work, and also approached local key informants and gatekeepers. Participants were invited by email or letter with all relevant details. All expenses incurred were paid by the project. Participants received compensation for their time to participate in the conversations: for regional conversations, we paid for accommodation, actual transportation costs, meals and a per diem of THB 400 per day. Participants in the online national conversations received THB 1000 per session.

In total, our Conversation events had 248 participants (179 individual attendees, some of whom attended more than one conversation event; see Suppl. Table 2 for meeting details and participant backgrounds). The regional conversations focused on regional issues and were held in Chiangmai, Khon Kaen, Hat Yai and Bangkok for participants residing in the northern, north-eastern, southern and central regions of Thailand respectively. For these, we worked with Civicnet Foundation, a community-based organisation with extensive experience in facilitating workshops in communities and inspiring change, to design and lead the activities. The first regional conversation ran over three full days (8 hours a day). In response to attendee feedback, the following three conversation events were shortened to 2.5 days. The national online conversations focused on national issues and were attended by people from all over the country. We conducted two sets of online conversations using Microsoft Teams: a ‘National Conversation’ and a Youth National Conversation’. Each online Conversation event included three 3-hour sessions with the same individuals (Suppl. Table 2). We invited 3-5 AMR stakeholders to join each community conversation, including key players in the NSP-AMR development, communication experts and AMR researchers. Agendas were modified to suit the context of each conversation event and informed by feedback from previous conversations; an example agenda is available online^26^.

In the conversations, we asked participants to generate as many ideas as possible, without going into too much detail about the practicalities of implementing these ideas. To narrow the scope, we focused on solutions aiming for more effective communication and engagement around AMR, and other tangible solutions actionable by participants themselves. In regional conversations, we asked participants to take into consideration their local health care system, and to create a concrete feasible one-year plan that they could implement in their communities. Suggestions that were not immediately actionable by participants, such as those around legislations or those to be actioned by other parties e.g. large pharmaceutical companies, World Health Organization (WHO), also emerged, but were not discussed in detail and are not reported here. After each conversation, the project team members reflected on the findings and summarized them. To produce the lists of key points, at the end of Phase II all findings and summary notes were analysed using an inductive approach of thematic analysis in interpreting the data into sub-themes, collapsed into bigger themes and linking into a core theme.

#### Phase III: Feedback to Planning/Community Conversation participants and other stakeholders

In this phase of the project, we presented findings from the Community Conversations to key AMR stakeholders through virtual and in-person meetings, conferences and reports (Table 1). We also invited relevant community leaders, change makers and ‘solution experts’ (e.g. communications experts) to some meetings. The findings and co-created solutions from this project were communicated to those involved in the writing of the NSP-AMR 2023-2027. Furthermore, we fed back our results to all participants using a printed booklet written in accessible language ^27^. Also included in the booklet were a series of original graphics for use in communication materials. They portray informal antibiotic sales in Thailand (Fig. 1) and scenes from the conversation events (Suppl. Fig 1).

**Figure 1:**
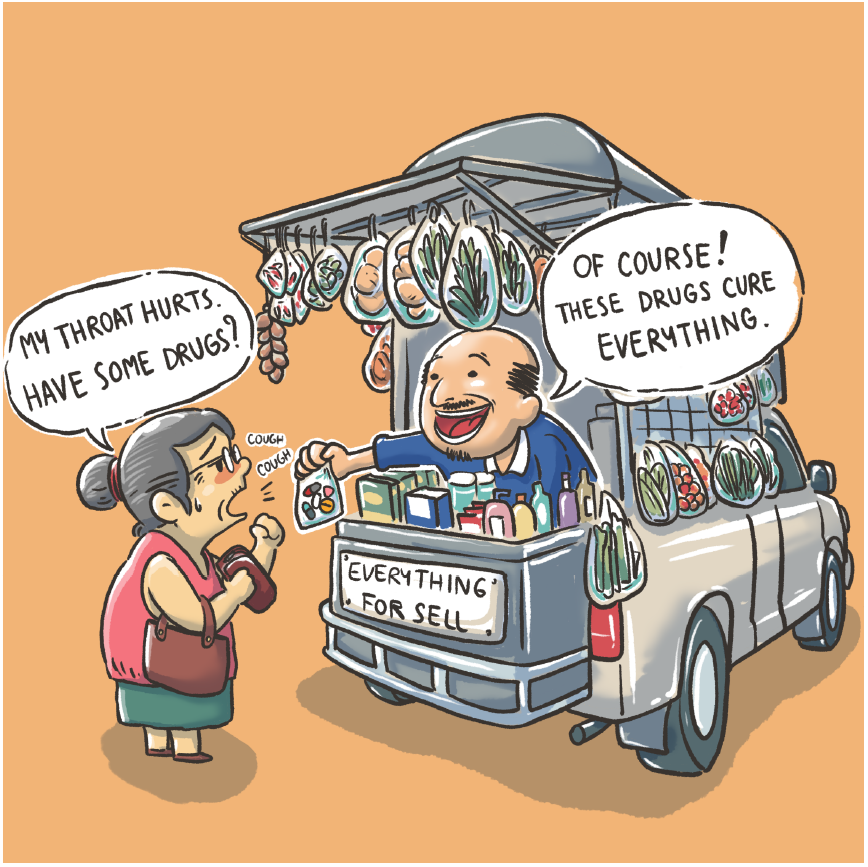
Cartoon portraying how easily available antibiotics are in Thailand. A woman complains that her throat hurts and asks for medicine, and a street vendor hands her a cocktail drug (‘*yaa chud*’) containing antibiotics from his cart, stating that these drugs cure everything (original art by team member KB).

### Problems and drivers for AMR in Thailand

Our project revealed four issues and drivers for AMR in Thailand.

#### 1) Misunderstandings around antimicrobials and AMR

Participants mentioned that many people in Thailand refer to antibiotics (in Thai 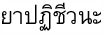 *ya patichiwana*) as ‘anti-inflammatories’ (in thai 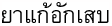 *ya kae ak-seb*). This often leads to a misunderstanding that antibiotics can treat muscle pain and inflammationn and hence be used regularly for aches, pains and fevers. Participants also mentioned that the words ‘antimicrobials’ (in Thai 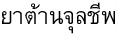 *ya tan jun la cheeb*) and ‘antibiotics’ in Thai are not easily understood, as they are formal words not used in everyday language. This is likely to be the reason why lay people and even healthcare professionals use other words (e.g. anti-inflammatories) to describe antibiotics and antimicrobials. Many participants did not know that antimicrobials also include antifungals, antiparasitics, and antivirals. This often leads to antibiotics being used for all types of illnesses, including common cold and COVID-19. Some people said that antibiotics are used often for prevention of illness or ‘just in case’.

#### 2) Limited reach and impact of communications and engagement around AMR

Thailand has taken actions to address AMR, including production of videos and printed materials, and organising the annual Antimicrobial Awareness Day/Week, held since 2013 ^28^. The Thailand Ministry of Public Health also launched public campaigns to warn people about the dangers of ‘*yaa chud*’ poly-pharmacy packs that contain antibiotics. However, many of the participants in the Community Conversations said that they were not aware of the activities. For many, the Community Conversations were the first time they heard of AMR (Suppl. Fig 1). When participants discussed the impact and limitations of existing materials, they thought that some of them were not suitable for the target group, and digital engagement strategies were not able to reach those without internet access or smartphones or older people. In-person events were only held in big cities and did not reach those who live in smaller towns and villages. Participants also mentioned that there is a ‘lack of faces’ or emotions in Thai AMR messaging. Lastly, due to a lack of evaluation it is not known to what extend these communication or engagement efforts achieved their target objectives.

#### 3) Social norms around taking and requesting antibiotics

All conversation events saw a lot of discussion around social norms. In Thailand, self-medication is practised broadly. When ill, many people buy medication at pharmacies or informal drug stores. Many people also mentioned that they themselves or someone they know share medications. Participants said many older people stop taking medications after their symptoms disappear, and often keep the unfinished medications for future use, either for themselves or for their families. Healthcare professionals mentioned that patients who see them in the clinic or hospital expect to be given some medication, even if the ailment is self-limiting. Doctors and pharmacists are also quick to prescribe antimicrobials for self-limiting ailments. Some participants referred to this an ‘addiction to antibiotics’. Others attributed it to lack of basic health literacy. Participants also mentioned that it is customary and polite, and even caring to ask friends and family members if they have taken any medication when they feel unwell.

#### 4) Availability of antimicrobials, limited monitoring and enforcement

Participants confirmed that antimicrobials can be easily obtained from doctors and other healthcare workers in government hospitals and clinics. They said that most antimicrobials can be dispensed by licensed pharmacists at pharmacies without a prescription from a qualified doctor. They added that pharmacies are widely available and antibiotics are inexpensive. Participants mentioned that generally there is limited enforcement of the regulations. Antibiotics are also sold illegally at grocery stores, mobile grocers and informal drug stores. Participants confirmed that it was not difficult to buy *‘yaa chud’*, which usually contains an antibiotic, a steroid, and an antipyretic, e.g. paracetamol, packed together in small unlabelled plastic bags. They are cheap to buy and easily available in village stores, but their quality is not assured. Participants also mentioned that antibiotics are used widely in poultry, fish farms, veterinary sector and agriculture. In some cases, animal grade antibiotics are repackaged and sold for human consumption.

### Co-created solutions: four locally actionable ‘building blocks’

In the course of the Conversation events, we collected many suggestions for potential solutions. Here we report only suggestions related to communications and engagement according to the scope of our project, which aimed to inform Strategy 5 (public knowledge and awareness of appropriate use of antimicrobials) of the NSP-AMR, and solutions actionable by the participants themselves. We outline four ‘building blocks’ of locally actionable solutions that participants suggested (detailed in Box 1):

- Messages around AMR should be clear and tailored to the target audience, and more frequent
- More initiatives to increase general health literacy
- Increased availability of AMR-related information at the local level
- Increased local leadership of AMR mitigation efforts

## Discussion

Thailand has made significant progress in improve its public health system,^29^ including efforts to mitigate the problem of AMR, such as establishing the Thailand National Strategic Plan on Antimicrobial Resistance 2017-2022 ^22 23^. The Thai NSP-AMR has six strategies ^23^. Our project aimed at informing Strategy 5, which at the time of the project focused on increasing public awareness of AMR; in the new NSP-AMR 2023-2027, Strategy 5 changed the focus to AMR literacy.

Our series of conversations confirmed some of the problem and drivers of AMR already known from other studies and the Thai national household surveys, for example that antimicrobials are still widely available including ‘*yaa chud*’ ^15 30-32^, and that there is still quite a lot of misunderstanding around antimicrobials ^16 17 23 31 32^. Our findings, in line with other studies, demonstrate the importance of community involvement and co-production in developing and implementing strategies to tackle AMR at a local level and promote community ownership ^31 33^.

In terms of solutions, participants suggested some changes to the current approaches to mitigate AMR. They recommended that communication messages regarding AMR should be concise, materials should be eye-catching and customized for the intended audience, there should be more programmes aimed at improving general health literacy, and local communities should have greater access to AMR-related information. This echoes what previously has been suggested by other studies: address the confusion between antibiotics and anti-inflammatories, explain the consequences of antibiotic overuse and misuse, e.g. for viral infections, include the importance of sanitation and hygiene ^32 34-37^, and tailor messages to the local context and specific misconceptions within a country ^38^.

The most striking finding was that many thought that local leadership should play a more prominent role in efforts to mitigate AMR, and mitigation strategies should take into consideration local culture. Participants suggested that in some places, AMR awareness and related projects should be combined with entertainment, whereas in other places, where community networks are strong (e.g. southern Thailand), projects should be embedded within existing networks and the primary health system, to address implementation gaps of the strategy.

Our project had several strengths and limitations (Table 2). One of the most notable outcomes of the geographical approach was that in some of the regional conversations, participants suggested low-hanging and locally actionable suggestions that they can initiate in their communities. In fact, a few informal local groups formed to organise local events after the conversation events (Box 2).

**Table 2:**
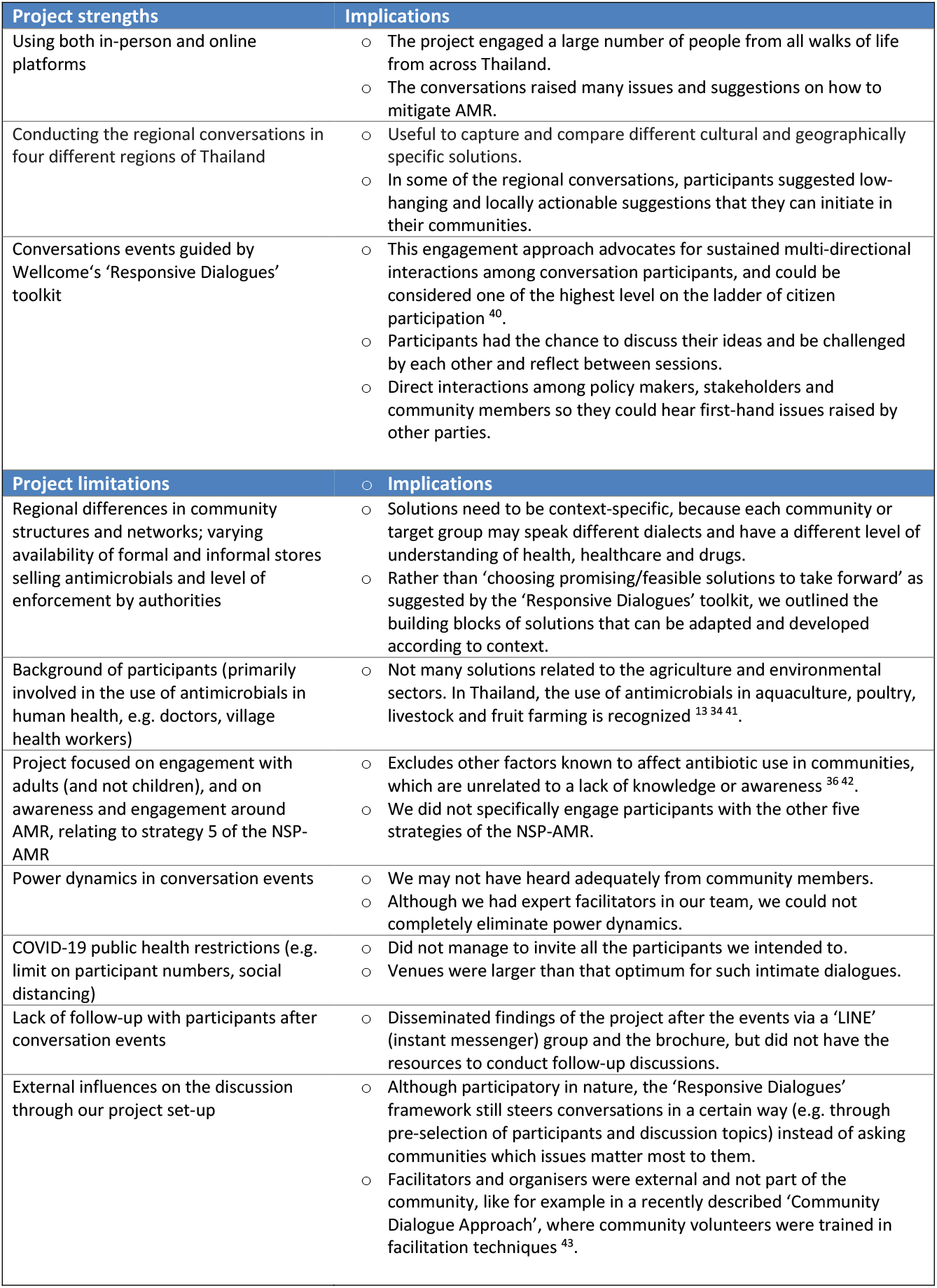
Strengths and limitations of the project and their potential implications.

| Project strengths | Implications |
| --- | --- |
| Using both in-person and online platforms | ○ The project engaged a large number of people from all walks of life from across Thailand.<br>○ The conversations raised many issues and suggestions on how to mitigate AMR. |
| Conducting the regional conversations in four different regions of Thailand | ○ Useful to capture and compare different cultural and geographically specific solutions.<br>○ In some of the regional conversations, participants suggested low-hanging and locally actionable suggestions that they can initiate in their communities. |
| Conversations events guided by Wellcome's 'Responsive Dialogues' toolkit | ○ This engagement approach advocates for sustained multi-directional interactions among conversation participants, and could be considered one of the highest level on the ladder of citizen participation<sup>40</sup>.<br>○ Participants had the chance to discuss their ideas and be challenged by each other and reflect between sessions.<br>○ Direct interactions among policy makers, stakeholders and community members so they could hear first-hand issues raised by other parties. |
| Project limitations | Implications |
| Regional differences in community structures and networks; varying availability of formal and informal stores selling antimicrobials and level of enforcement by authorities | ○ Solutions need to be context-specific, because each community or target group may speak different dialects and have a different level of understanding of health, healthcare and drugs.<br>○ Rather than 'choosing promising/feasible solutions to take forward' as suggested by the 'Responsive Dialogues' toolkit, we outlined the building blocks of solutions that can be adapted and developed according to context. |
| Background of participants (primarily involved in the use of antimicrobials in human health, e.g. doctors, village health workers) | ○ Not many solutions related to the agriculture and environmental sectors. In Thailand, the use of antimicrobials in aquaculture, poultry, livestock and fruit farming is recognized<sup>13 34 41</sup>. |
| Project focused on engagement with adults (and not children), and on awareness and engagement around AMR, relating to strategy 5 of the NSP-AMR | ○ Excludes other factors known to affect antibiotic use in communities, which are unrelated to a lack of knowledge or awareness<sup>36 42</sup>.<br>○ We did not specifically engage participants with the other five strategies of the NSP-AMR. |
| Power dynamics in conversation events | ○ We may not have heard adequately from community members.<br>○ Although we had expert facilitators in our team, we could not completely eliminate power dynamics. |
| COVID-19 public health restrictions (e.g. limit on participant numbers, social distancing) | ○ Did not manage to invite all the participants we intended to.<br>○ Venues were larger than that optimum for such intimate dialogues. |
| Lack of follow-up with participants after conversation events | ○ Disseminated findings of the project after the events via a 'LINE' (instant messenger) group and the brochure, but did not have the resources to conduct follow-up discussions. |
| External influences on the discussion through our project set-up | ○ Although participatory in nature, the 'Responsive Dialogues' framework still steers conversations in a certain way (e.g. through pre-selection of participants and discussion topics) instead of asking communities which issues matter most to them.<br>○ Facilitators and organisers were external and not part of the community, like for example in a recently described 'Community Dialogue Approach', where community volunteers were trained in facilitation techniques<sup>43</sup>. |

## Conclusion

The conversations using the ‘Responsive Dialogues’ approach unearthed many local issues and produced four ‘building blocks’ of locally actionable solutions. Our findings will be relevant to those who would like to involve communities and other stakeholders in their work, those who create tools or interventions for uptake by communities or local authorities, organisations who produce communication materials for increasing AMR or health literacy, and those looking for tangible policy solutions at a local level.

## Abbreviations

AMR: Antimicrobial resistance
NSP-AMR: Thailand National Strategic Plan on Antimicrobial Resistance

## Data availability

Data underlying the paper may be requested from the Mahidol-Oxford Tropical Medicine Data Access Committee.

## Author contributions

PYC wrote the first draft of the paper and raised the funding. PYC, AO, DL, CT, NKA and TP designed the project. TP was the overall project manager. TP, RP, SK, CT, KS, NKA, RP and KB facilitated the conversation events. BN led the analysis of the findings and evaluation. NKA, DL, NS and SP provided input and guidance on the alignment of the project with the Thailand National Strategic Plan on Antimicrobial Resistance. KB created the illustrations. All authors reviewed and approved the final version of the protocol. PYC is the principal investigator and the guarantor of the paper.

## Acknowledgements

We would like to thank all of the participants in our conversation events for their insightful contributions, and for taking the time to attend our sessions. We thank members of the Bangkok Health Research Ethics Interest Group for their input in the development of the project. We also thank Lucy McDowell, Zaichen Mallace-Lu and Jo Zaremba for their support and intellectual input throughout the duration of the project.

## Box 1

The four co-created ‘building blocks’ of solutions to tackle AMR and detailed recommendations from the conversation events

**Messages around AMR should be clear and tailored to the target audience**

⇒ Tailor information materials according to target audience.
⇒ Adapt messages according to context and use the local dialect, or illustrations suitable for the target group.
⇒ Messaging should use channels preferred by the target group (e.g. with youth groups, social media platforms are most popular).
⇒ Materials should preferably be informed by communications and behaviour change research and tested with the target group before roll-out.
⇒ Embed monitoring and evaluation in communication and engagement initiatives. In some communities, it may be useful to engage community influencers or leaders to pass on the knowledge.

**More initiatives to increase general health literacy**

⇒ Instead of only focusing messaging on antimicrobials and AMR, there should be a move to increase holistic health literacy, which includes sanitation, nutrition and wellness.
⇒ Incorporate AMR and health literacy in the school curriculum and informal learning centres for adults.

**Increased availability of AMR-related information at the local level**

⇒ Participants at the local level, such as village health volunteers and healthcare staff at the primary care level, would like AMR-related information to be ‘returned to the community’. These include local data on usage of antimicrobials, deaths due to drug resistant infections and stories at local level. This way, they can make better informed decisions in relation to antimicrobial usage and provide information to their communities.

**Increased local ownership of AMR mitigation efforts**

⇒ Support shared leadership and increased local leadership in mitigating the problem of AMR.
⇒ Community leaders are best placed to create awareness and share knowledge on health and AMR because they know how to engage with their own community.
⇒ Local level administration is more permanent than higher level administration and politicians, and there are shorter command chains to implement any activities or programmes.
⇒ Local leaders can establish their own AMR task force and community surveillance on unauthorised sales of antimicrobials. This concept is called *bo-worn* in Thai, which means sublime, heavenly or great. *Bo-worn* consists of three parts, *bo, wo and ro* which is short *for baan* (house or for this context means community or group); *wat* (temple, mosque or church which are places of warship) and *rong-rean* (school and educational institutions). This concept has been used widely to promote and strengthen local networks to address local issues in a sustainable way^**44 45**^.
⇒ Participants said they themselves will find opportunities to raise awareness of AMR in the communities. These include local talks, information sessions and in-person, one-on-one ‘heart-to-heart’ conversations (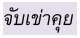*(jap khao kooi)”*.
⇒ Solutions should take into consideration the culture and preferences of engagement of each region:
  - Northern region: folk story telling
  - North-eastern region: fun and light-hearted activities
  - Southern region: family or community-based activities
  - Central region and other urban areas: social media.

## Box 2

List of informal local groups and events that participants organised after attending the conversation events.

- Northern region: Khun Yuam Hospital, Mae Hong Son province, in collaboration with informal school to teach AMR to parents and older people at “Elderly School” in March 2022.
- Southern region: The village health volunteers (VHV) from Pattani and Pattalung Province brought AMR as topic to their VHV monthly meeting in April 2022.
- Central region: The village health volunteer leader from the Klong Toey slum requested an AMR expert to provide training on basic AMR knowledge for village health volunteers in Bangkok’s Klong Toey slum in July 2022.

