## Supplementary files for "Embedding community and public voices in co-created solutions to mitigate antimicrobial resistance (AMR) in Thailand using the ‘Responsive Dialogues’ public engagement framework"

**Supplementary Table 1.** Extended Thai antimicrobial resistance stakeholder map.

| *Stakeholder group* | *Stakeholder details* |
| --- | --- |
| Government organisations | - National, regional and local policy makers and government officers - Regulators - Authorities:   - Ministry of Public Health (MoPH), Thailand   - Ministry of Agriculture and Cooperatives   - Ministry of Science and Technology   - Ministry of National Resource and Environment Health Education Division, MoPH   - Ministry of Education   - Government Office of the Non-formal and Informal Education   - Ministry of Labour   - The Government’s Public Relations Department (กรมประชาสัมพันธ์)   - Food and Drug Administration, MoPH   - National Antimicrobial Resistance Surveillance Center, Thailand   - Bamrasnaradura Infectious Disease Institute, DDC, MoPH   - Division of Epidemiology, Department of Disease Control, MoPH   - Department of Medical Service, MoPH   - Bureau of Health Administration, MoPH   - Bureau Sanatorium and Healing Arts, Department of Health Service Support, MoPH   - Division of Primary Health Care, Department of Health Service Support, MoPH   - Bureau of Food and Water Sanitation, Department of Health, MoPH   - Bureau of Quality and Safety of Food, Department of Medical Sciences, MoPH   - Bureau of Drug Control, Food and Drug Admistration, MoPH   - Bureau of Emerging Infectious Diseases, Department of Disease Control, MoPH   - Bureau of Food Control, Food and Drug Administration, MoPH   - Technical and Policy Administration Division, Food and Drug Administration, MoPH   - Public and Consumer Affairs Advertisement Control Division, Food and Drug Administration, MoPH - Doctors and nurses from the Health Promotional Hospital (รพสต. Primary Care Unit (PCU), or Anamai อนามัย) of that district/sub-district - International agencies - Healthcare Accreditation Institute (Public Organization) - National Health Security Office - National Health Commission Office |
| Public sector/Non-governmental organizations/Civil society organizations | - Civicnet Foundation - Folk Doctor Foundation - World Animal Protection - Greenpeace, Thailand - The College of Pharmaceutical and Health Consumer Protection of Thailand (CPHCP) - Community organisations - Community engagement teams - Mass media agencies - Chaladsue magazine, foundation for consumers - Patient associations e.g. Thai Caregivers Association, Thai Association Diabetes Educator - Citizen Associations, e.g. Thailand Active Citizen, Thailand HelpAge, Thailand Be Health - Consumer Associations, e.g. Foundation of Consumers, The Association of Confederation of Consumer Organisation, Thailand (ACCOT) - Animal Rights associations e.g. Thai Animal Guardians Association, World Animal Protection |
| Academic/researchers | - Lecturers and students in the following disciplines: pharmacy, healthcare, medicine, global health, microbiology, social science, public relations, etc. - Researchers - Research networks - AMR-focused international networks - Communicators   **For example:**   - Mahidol-Oxford Tropical Medicine Research Unit (MORU), Shoklo Malaria Research Unit (SMRU) - Faculty of Tropical Medicine, Mahidol University - Faculty of Pharmacy, Mahidol University - Faculty of Social Sciences & Humanities Mahidol University and Antimicrobials in Society (AMIS) - Drug System Monitoring & Development Center (DMD), Faculty of Pharmaceutical Sciences, Chulalongkorn University - Graduate School of Communication Arts and Management Innovation, National Institute of Development Administration (NIDA) - RTG-WHO Country Cooperation Strategy on AMR program 2017-2021 (Food and Drug Administration and International Health Policy Program) - Health Systems Research Institute - International Health Policy programme - International organizations supporting AMR research or partnership with Thai partners, e.g. World Health Organization, Food and Agriculture Organization of the United Nations, United States Agency for International Development, Centers for Disease Control and Prevention, Wellcome - ReAct - Sonar-Global - A Clinically Oriented Antimicrobial Resistance Network (ACORN) |
| Private sector | - Suppliers: manufacturers, importers, distributors, generic companies - Producers (e.g. food) - Pharmaceutical companies (international and local) - Private clinics and health providers - Community pharmacy - Retailers, online retailers - Grocery shops - Other sellers/retailers |
| Healthcare sector | **Setting:**   - Hospitals 1. Super tertiary care (university hospitals) 2. Tertiary care (medical center and regional hospital [over 500 beds]) 3. Secondary care (general hospital [120 –500 beds] and community hospitals [10 – 120 beds] 4. Primary care (health promotion hospital) - Health centers - Public Health provincial office - Pharmacy/drug stores - Private clinics - Pharmacy association   **Person:**   - Doctors - Sub-district medical practitioner - Nurses - Primary Care Unit (nurses, public health officers) - Healthcare workers - Community healthcare volunteers - Thai village health volunteers (อสม) - Pharmacists - Dentists - Traditional healers - Massage practitioners - Public health officers (นักวิชาการสาธารณสุข) |
| 'One Health’ sectors | - Farmers (terrestrial and aquaculture) - Consumers - Vets - Animal husbandry - Animal hospitals - Animal pharmacies - Pet shops and animal shelters - Agricultural shops - Markets and retailers - Feed mills - One Health Drivers of Antibacterial Resistance in Thailand (OH-DART) - Ministry of Agriculture and Cooperatives - Department of Fisher - Department of Agriculture - Department of Agriculture extension - Animal Feed and Veterinary Products Control, Department of Livestock Development - National Institute of Animal Health, Department of livestock Development - Bureau of Quality Control of Livestock Products Dept of Livestock Development |
| Members of the public | - User/consumer (farmers, children, parents, adolescent, working-age) - Teachers and educators:   - formal education   - non-formal/adult education - Parents and extended families - Communities and networks - Head of community, Head of villagers, village leaders - Youth groups - Community Advisory Boards - Youth Advisory Boards - Actors/actresses - Monks, temple staff/speaker |
| Media and solution experts | - International and local TV, e.g. Thai PBS, Thairath, Amarin, MCOT HD, Nation TV - Print media - Journalists - Social media experts - Social media influencers - Artists - Animation experts - Science communicators - Reporters |

**Supplementary Table 2:** Additional information about all Conversation events, including meeting length, number of participants, demographics and professional backgrounds.

| Activity | Date | Meeting length | Number of participants^1^ | Number of men/women/ people who identified as ‘other’ | Age range | Participant background^2^ | Province/ region |
| --- | --- | --- | --- | --- | --- | --- | --- |
| PHASE I | | | | | | | |
| Planning Conversation 1  (in-person) | 25^th^ Nov 2020 | 0.5 days | 21 | M=5  F=11 | 28 - 65 | - AMR government officers - AMR researchers and experts - Healthcare providers - Non-governmental organizations (Greenpeace Thailand, World Animal Protection) - Communication and solution experts | Bangkok |
| Planning Conversation 2 (online during COVID-19 restrictions) | 7^th^ Jan 2021 | 3 hours | 20 | M=4  F=16 | 30 - 67 | As above | Thailand |
| Planning Conversation 2  (in-person) | 31^st^ March – 1^st^ April 2021 | 2 days | 26 | M=11  F= 15 | 28-71 | As above | Bangkok |
| PHASE 2 | | | | | | | |
| National Conversation (online) | 6^th^ May, 2^nd^ June and 1^st^ July 2021 | 3 hours | 18 | M=7  F= 11 | 20-69 | - Healthcare providers, e.g. medical doctors, pharmacists - Business owners - Students - Freelancers - Housewife - Senior editor in health magazine - Science communicator | Thailand |
| Youth National Conversation (online) | 3^rd^, 10^th^, and 17^th^ Nov 2021 | 3 hours | 30 | M=12  F=17  Other =1 | 18 -24 | - High school students (grade 12) - University students (e.g. public health, pharmacy, public administration program, commerce and accountancy, social administration, education, engineering, anthropology) - Employees (e.g. office workers, engineer, information technology, case manager ) - Unemployed | Thailand |
| Northeastern conversation | 14^th^ –16^th^ Dec 2021 | 3 days | 26 | M= 9  F=17 | 31- 70 | - Healthcare providers, e.g. pharmacists, dentist, nurses, public health officers, village health volunteers, - Farmers - Grocery owner - Community leaders - Local administration organization staff | Khon Kaen |
| Northern conversation | 21^st^ –23^rd^ Feb 2022 | 2.5 days | 24 | M=4  F=20 | 30-64 | - Healthcare providers, e.g. pharmacists, nurses, public health officers, village health volunteers - Member of consumer Northern network - Teachers - Farmers | Chiangmai |
| Southern conversation | 29^th^ – 31^st^ March 2022 | 2.5 days | 27 | M= 8  F= 19 | 28-70 | Healthcare providers, e.g. medical doctor, pharmacists, nurses,   - Public health officers, village health volunteers, - Farmers - Volunteer teacher - Civil Society leaders - Local administration organization staff - Member of Thailand Consumers Council | Hat Yai |
| Central conversation | 18^th^ –20^th^ May 2022 | 2.5 days | 23 | M=11  F= 12 | 19-74 | - Healthcare providers, e.g. medical doctor, pharmacists, nurses, - Public health officers, village health volunteers in slum - NGOs, social campaigners - Radio broadcasters - Health communicator - Marketing specialist - Leader on organic farm | Bangkok |
| PHASE 3 | | | | | | | |
| Feedback session 1 | 17^th^ June 2022 | 1 day | 18 | M=9  F=9 | 34-74 | Same participants from Planning Conversations, including representative participants from 4 region | Bangkok |
| Feedback session 2 | 8^th^ July 2022 | 1 day | 15 | M=6  F=9 | 34-74 | Same participants from Planning Conversations including representative participants from 4 region | Bangkok |
| Total |  |  | **248** | **M= 86**  **F= 156**  **Other = 1** |  |  |  |
| Total number of individual participants^3^ |  |  | **179** |  |  |  |  |
| *Notes*:   ^1^ This number excludes the project team, Civicnet facilitators, and members of the MORU Bioethics and Engagement team.  ^2^ Some participants held multiple identities, e.g. some participants in the community conversations worked as farmers, but also had responsibilities as community leader.  ^3^ For the Planning Conversations, regional conversations and feedback sessions, the same participants were invited to all events, meaning that many of them attended two or more events. We chose to include the total number of participants in each conversation to give an idea of the group size for each event, and to include the total number of individual participants at the end to show how many people engaged with the project. | | | | | | | |

**Suppl. Figure 1**. Cartoon-style drawings capturing the atmosphere during the community conversation events. A) Participants from diverse backgrounds (villager, policy maker, farmer, village health volunteer = VHV) having a conversation about AMR. B) The regional in-person conversations started with observing mindfulness each day. C) Participants are surprised to learn about the seriousness of AMR.

| 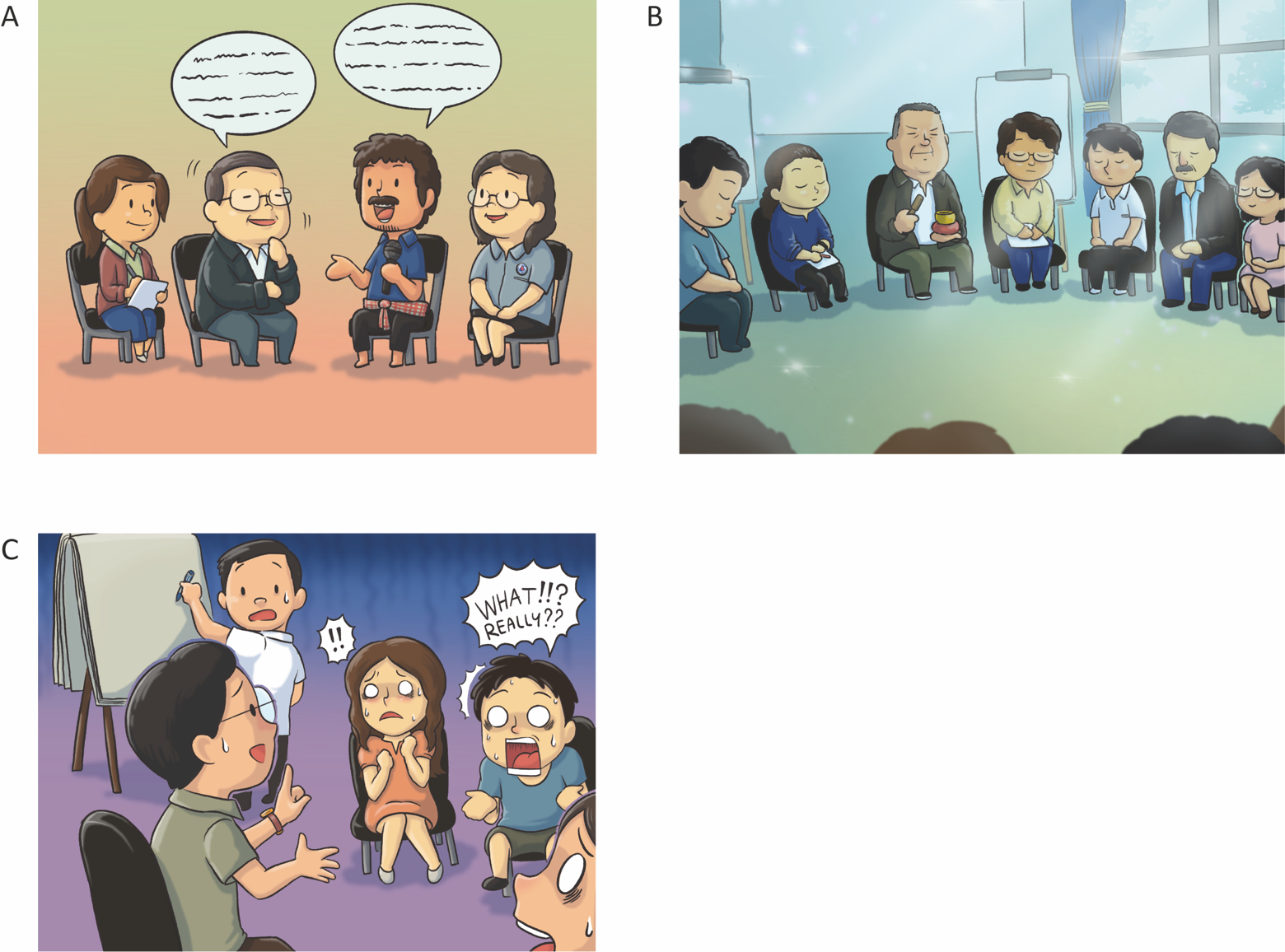 |
| --- |
